# To have or not to have (knee arthroplasty) two years after a simple home-based exercise program in patients with severe knee osteoarthritis eligible for knee arthroplasty: A 2-year exploratory follow-up of the QUADX-1 randomized trial

**DOI:** 10.64898/2026.04.30.26351578

**Authors:** Thomas Bandholm, Birk Grønfeldt, Rasmus Skov Husted, Emma Stenholm Koch, Anders Troelsen, Helle Gybel Juhl-Larsen, Kristian Thorborg

## Abstract

**Background and purpose:** In the QUADX-1 trial, we randomized 140 patients with severe knee osteoarthritis (OA) eligible for a knee arthroplasty to home-based exercise for 12 weeks. Seventy-nine (68%) of the 117 patients, who completed the exercise intervention, postponed surgery. Here, we report how many patients, who completed the 12-week exercise intervention, had received a knee arthroplasty at 2 years and describe their initial exercise response.

**Methods:** From the QUADX-1 trial, we had the following: isometric knee-extensor strength, Oxford Knee Score (OKS), Knee Osteoarthritis Outcome Score (KOOS), average knee pain last week (0-10 numeric rating scale [NRS]), 6-minute walk test, stair climbing test, and self-reported exercise behaviour.

**Results:** At the 2-year follow-up, 50 (43%) of the 117 patients had received a knee arthroplasty (KA group) and 67 (57%) had not (NO-KA group). Compared with the KA group, the NO-KA group had less severe radiographic OA at baseline (KL grade 4: 38% vs 55%) and showed greater— and often clinically relevant—improvements after the 12-week exercise intervention, including knee pain (-2.1 vs -0.1 NRS points), OKS (+6.9 vs +0.5 points), and KOOS ADL (+13.9 vs +1.3 points).

**Conclusion:** Two years after completing the initial 12-week QUADX-1 exercise intervention, more than half the cohort had not received a knee arthroplasty despite initially being considered eligible. Those who had not received a knee arthroplasty at two years had less severe radiographic OA at baseline and generally responded better to 12-week exercise two years earlier, compared to those who had.

**ClinicalTrials.gov-ID:** NCT02931058.

## INTRODUCTION

The NICE and OARSI guidelines for the treatment of knee osteoarthritis state that knee arthroplasty should be considered for patients with severe symptoms that substantially impact quality despite non-surgical management (1,2). Finding the right patients to have surgery at the right time is challenging, as the indication for a knee arthroplasty (KA) is not clear cut, and coordination between non-surgical and surgical management is often suboptimal (3,4).

In the QUADX-1 trial (5), we tried a simple model of coordinated non-surgical (exercise) and surgical care of patients eligible for a KA to ensure coordination and enhance surgical decision making. The QUADX-1 trial was a randomized dose-response trial that used a minimal treatment approach to home-based exercise (a program of only 1 exercise [seated knee extensions]) (5). We saw no signs of an exercise-dose outcome-response relationship, but found that 68% of all participants decided not to have surgery after the 12 weeks (5). Similar findings have been reported by others after more comprehensive exercise interventions (6–9). The question is how long such an exercise effect may last, especially one induced by a program consisting of only one exercise (5).

Although 68% of all patients who completed the initial QUADX-1 exercise intervention chose to initially postpone surgery, peers have asked: “Promising, but aren’t we just kicking the can down the road? For how long does this last?” This skepticism in the long-term effect was also reflected in the surgeon interviews when asked about the QUADX-1 intervention being a non-surgical component in a coordinated care pathway (10). In this 2-year follow-up, we address this relevant question and skepticism by reporting the 2-year surgical status. We report how many patients, who completed the 12-week exercise intervention, had received a KA at 2 years and describe their initial exercise response.

## METHODS

### Study design

In the QUADX-1 trial (5) we randomized patients with severe knee OA eligible for KA to a program of *one* home-based, knee-extensor exercise for 12 weeks with dosages of 2, 4, or 6 sessions per week. We saw few between-dosage differences across outcomes, but 79 (68%) of the 117 patients, who completed the exercise intervention, postponed surgery, unrelated to the exercise dosage (trial not powered for this outcome and comparison) (5). Here we report a 2-year follow-up with a focus on surgical (KA) status and responses to the initial 12-week exercise intervention in those who completed it. The QUADX-1 trial was pre-registered (NCT02931058) and approved by the Ethics Committee of the Capital Region, Denmark (H-16025136) as well as the Danish Data Protection Agency (2012-58-0004), including the 2-year follow-up. Written informed consent was obtained from all participants. The 2-year follow-up of the QUADX-1 trial was not pre-defined (11), which is why we consider this an exploratory trial follow-up. AI (ChatGPT) was used for proofreading and grammar. The AI output was controlled by a human (specialist).

### Participants

Patients potentially eligible for trial participation were recruited at the surgical outpatient clinic. The patients were referred from their general practitioner for surgical evaluation by an orthopedic surgeon specialized in KA. The patients were told that if they agreed to participate, they would complete the home-based exercise intervention and afterwards their need for surgery would be re-evaluated in a shared decision-making process between the patient and orthopedic surgeon (e.g., continue with exercise or schedule surgery).

The inclusion criteria were: eligible for KA due to knee OA (assessed by an orthopedic surgeon), radiographically verified knee OA with Kellgren-Lawrence classification ≥ 2, average knee pain ≥ 3 (Numeric Rating Scale (NRS)) in the last week, eligible for home-based knee-extensor exercise, age ≥ 45 years, resident in one of three municipalities involved in the trial (Copenhagen, Hvidovre or Brøndby) and able to speak and understand Danish. The exclusion criteria were: exercise being contraindicated, neurological disorder, diagnosed systemic disease (American Society of Anesthesiologists’ physical status classification score ≥ 4), terminal illness, severe bone deformity demanding use of non-standard implants, or a greater weekly alcohol consumption than the national recommendation.

### Patient and public involvement

Patients were involved and gave feedback during pilot testing of QUADX-1 trial procedures.

### Interventions

Detailed intervention descriptions and all related intervention material can be found in the open access trial protocol (11) or primary trial report (5), including a link to an instructional video (http://bit.ly/2EV7NrF). Briefly, following baseline assessment, the patients were referred to a physiotherapist at their local municipality rehabilitation setting. Here the patients were instructed how to perform a single knee-extensor exercise at home. The patients were randomized to one of three weekly exercise dosages: two, four or six sessions per week for twelve weeks, using 12 Repetition Maximum (RM) intensity sets to be performed at home. They had control visits with the municipality physical therapist after 4 and 8 weeks, at which they could ask questions related to the exercise.

### Variables and outcome measures

Outcomes for this 2-year follow-up were collected at three time points; (1) at baseline (before the start of the exercise intervention), (2) after twelve weeks of home-based exercise, and (3) 2 years after the end of the 12 weeks of exercise (in those who did not have a KA within 2 years) or 2 years after their day of surgery (in those who had surgery within the 2 years after the 12 weeks of exercise). For time points 1 and 2 (before and after the 12-week exercise intervention), the outcomes used to describe the response to the intervention were: knee pain (0-10 numerical rating scale [NRS] (12) and 0-100 KOOS (13) pain subscale, 100 best), self-reported; Activities of Daily Living, other Symptoms, knee-related Quality of Life; and Sport and Recreation function (KOOS: ADL, Symptoms, QoL, Sport/Recreation 0-100 point subscales, respectively, 100 best), self-reported function and pain (Oxford Knee Score (OKS) (14) 0-48 points, 48 best), performance-based function (6-minute walk (15) and stair climbing tests (16)), and knee-muscle (quadriceps) function (isometric knee-extension strength at 60 deg flexion (17); QUADX-1 primary outcome).

These self-reported and performance-based outcomes were selected to reflect constructs typically affected by OA and constructs commonly targeted with interventions. A detailed description of outcomes can be found in the published trial protocol (11). At time point 3, two years after the intervention or two years after the KA, questionnaires were sent to all participants electronically using a secure web application (RedCap (18)) or physically using postal mail, depending on participant preference. Surgical status (KA or not) was extracted from the electronic patient records. The questionnaires were: KOOS, OKS, and a QUADX-1 questionnaire containing questions on use of the elastic exercise band. The questions were: (1) “Have you exercised your (OA) knee using the elastic exercise band since the intervention stopped two years ago?” with response categories of “Yes”, “Yes, as needed”, “No”, and “No, I have participated in other knee-focused exercise”, (2) “How often have you exercised your (OA) knee using the elastic exercise band” (if answering yes to (1)) with response categories of “Monthly” and “Weekly”.

### Blinding

All outcome assessors and the data analysts were blinded to the exercise group allocation for the QUADX-1 trial. All data analyses for the 2-year follow-up were unblinded, meaning that surgical status at two years was used as a grouping variable and known to the data analyst.

### Sample size and power considerations

We did not estimate a sample size specifically for this 2-year follow-up, so all analyses are considered exploratory. Sample size justification for the QUADX-1 trial can be found in the published trial protocol (11).

### Statistical analyses

Depending on the distribution of the variables, patient characteristics are presented either as numbers with percentages (%) for categorical variables, and as means with standard deviations, or as medians with interquartile ranges (IQR) for continuous variables. In exploratory analyses, we investigated the between-group difference in change in the above-listed outcomes from baseline to the 12-week follow-up using two-sample t-tests (Table 2). For descriptive long-term analyses, KOOS and OKS values at the third time point were presented separately for participants with complete 2-year questionnaire data across the relevant time points (Table 3). Within-group changes from baseline to the third time point and from the 12-week follow-up to the third time point were investigated using paired t-tests (Table 3). Level of statistical significance was set at 0.05 and all p-values were corrected for multiple testing using the Bonferroni correction (multiplying the p-value by 9 tests performed for results presented in Table 2, and by 6 tests performed for results presented in Table 3). Data management was conducted using the SAS 9.4 software package for Windows (SAS Institute, Cary, NC, USA). R version 4.1.0 (The R Foundation for Statistical Computing, Vienna, Austria) was used for statistical analyses.

## RESULTS

Of the 117 patients who completed the 12-week QUADX-1 exercise intervention, 50 (43%) patients had received a KA (KA group) at the 2-year follow-up and 67 (57%) had not (NO-KA group). Thirty-two of the 117 patients chose to have a KA immediately after the QUADX-1 intervention (5), 8 additional patients had a KA within the first year after the QUADX-1 intervention, and 10 patients had a KA between the first and the second year after the QUADX-1 intervention. At the 2-year follow-up, 31 of 51 questionnaire responders in the NO-KA group (61%) reported having exercised their affected knee with the elastic band at some point after the QUADX-1 intervention; 13 (26%) reported regular use and 18 (35%) reported using it as needed. Fourteen patients (28%) had not used the elastic band, and six (12%) reported participating in other types of knee-focused exercise instead.

The NO-KA group had more patients with a Kellgren-Lawrence (KL) score of 2 and 3 compared to the KA group at baseline (Table 1). Other than the KL-scores, the groups were generally comparable across outcomes at baseline with differences below clinical relevance (Table 1 and 2). Following the 12-week exercise intervention, the NO-KA group generally demonstrated greater and often clinically relevant improvements across outcomes, compared to the KA group (Table 2). At the 2-year follow-up, among participants with complete 2-year questionnaire data, the respondents in the NO-KA group had better OKS and KOOS subscale scores than at baseline (P<0.05, Table 3), but generally not better than after the 12-week exercise intervention. In contrast, respondents in the KA group had markedly better OKS and KOOS scores 2 years after their KA than both at baseline and at 12-week follow-up (P<0.05, Table 3). No formal between-group statistical comparisons were performed for Table 3, and the between-group differences at the third time point should therefore be interpreted descriptively only.

**Table 1:** Baseline characteristics of the participants who completed the 12-week QUADX-1 exercise intervention stratified by 2-year surgical status and total cohort.

|  |  |  |  |
| --- | --- | --- | --- |
| Variable | NO-KA group<br>(N=67) | KA group<br>(N=50) | Total cohort<br>(N=117) |
| Female sex, N (%) | 34 (50.7) | 28 (56) | 62 (53) |
| Kellgren-Lawrence score, n (%) |  |  |  |
| Grade 2 | 11 (16.7) | 4 (8.2) | 15 (13) |
| Grade 3 | 30 (45.5) | 18 (36.7) | 28 (41.7) |
| Grade 4 | 25 (37.9) | 27 (55.1) | 52 (45.2) |
| Age, yrs; mean (SD) | 66.4 (11.0) | 67.6 (8.5) | 66.9 (10.0) |
| Body mass index; mean (SD) | 32.1 (7.2) | 31.3 (5.0) | 31.8 (6.4) |
| Abbreviations: KA=knee arthroplasty; N=number; SD=standard deviation. |  |  |  |

**Table 2:** Change from baseline to 12-week follow-up stratified by 2-year surgical status.

|  |  |  |  |  |  |  |  |
| --- | --- | --- | --- | --- | --- | --- | --- |
| Variable | NO-KA group<br>(N=67) |  |  | KA group<br>(N=50) |  |  | P-value* |
|  | Baseline | 12-week FU | Δ<sub>12-week FU-baseline</sub> | Baseline | 12-week FU | Δ<sub>12-week FU-baseline</sub> |  |
|  | Mean (SD) | Mean (SD) | Mean (95% CI) | Mean (SD) | Mean (SD) | Mean (95% CI) |  |
| Current knee pain, NRS points | 2.5 (2.3) | 1.7 (2.2) | -0.8 (-1.4, -0.3) | 1.8 (1.9) | 2.1 (2.4) | 0.3 (-0.5, 1.0) | 0.094 |
| Average knee pain, NRS points | 5.7 (1.7) | 3.7 (2.1) | -2.1 (-2.6, -1.6) | 5.8 (1.4) | 5.8 (1.8) | -0.1 (-0.6, 0.5) | <0.001 |
| OKS, points | 25.7 (7.8) | 32.6 (9.1) | 6.9 (5.2, 8.7) | 24.4 (7.4) | 24.9 (8.0) | 0.5 (-1.2, 2.2) | <0.001 |
| KOOS |  |  |  |  |  |  |  |
| Pain, KOOS points | 51.7 (17.4) | 67.1 (18.8) | 15.4 (12.2, 18.7) | 48.6 (15.4) | 50.2 (17.6) | 1.7 (-2.6, 6.0) | <0.001 |
| ADL, KOOS points | 57.5 (19.5) | 71.4 (20.2) | 13.9 (10.6, 17.3) | 54.5 (15.8) | 55.8 (17.5) | 1.3 (-3.5, 5.7) | <0.001 |
| QoL, KOOS points | 36.1 (17.0) | 47.9 (21.6) | 11.8 (7.1, 16.5) | 29.0 (14.4) | 31.9 (16.7) | 2.9 (-1.7, 7.5) | 0.084 |
| Symptoms, KOOS points | 55.8 (19.1) | 69.7 (19.7) | 13.9 (10.1, 17.8) | 55.5 (18.9) | 57.0 (22.1) | 1.5 (-2.6, 5.6) | <0.001 |
| Sport, KOOS points | 22.3 (22.8) | 35.4 (26.0) | 13.2 (7.9, 18.4) | 19.9 (18.5) | 22.0 (21.8) | 2.1 (-3.3, 7.5) | 0.047 |
| Knee-extension strength, Nm/kg | 1.3 (0.6) | 1.4 (0.6) | 0.2 (0.1, 0.2) | 1.2 (0.5) | 1.3 (0.5) | 0.1 (0.1, 0.2) | 1.000 |
| 6-minute walk test, m | 428.9 (100.6) | 460.6 (105.5) | 34.4 (22.6, 46.2) | 420.3 (92.0) | 432.9 (93.2) | 7.4 (-8.7, 23.7) | 0.100 |
| Stair climb test ascend, sec | 9.4 (4.9) | 8.1 (3.4) | -1.4 (-2.0, -0.8) | 8.5 (3.5) | 8.4 (4.1) | 0.2 (-0.3, 0.7) | 0.002 |
| Stair climb test descend, sec | 10.5 (6.7) | 8.4 (4.8) | -2.2 (-3.1, -1.4) | 9.3 (5.5) | 9.0 (4.8) | 0.2 (-0.5, 1.0) | <0.001 |
| Note: Current knee pain = How painful is your knee right now? Average knee pain = How painful has your knee been on average over the past two weeks?<br>Abbreviations: ADL=Activities of Daily Living; 95% CI=95% confidence interval; Diff=Difference; FU=follow up; KA=knee arthroplasty; KOOS=Knee Osteoarthritis Outcome Score (0-100 (best)); m=meters; 6MWT=6-minute walk test; N=number; Nm=Newton meter; NRS=Numeric Rating Scale; OKS=Oxford Knee Score (0-48 (best)); QoL=Quality of Life; SD=standard deviation; sec=seconds; SCT=Stair Climb Test; 12-w FU=12-week follow-up assessment. *Bonferroni-corrected P-values for between-group differences. |  |  |  |  |  |  |  |

**Table 3:** Outcome values across time points stratified by 2-year surgical status in participants with complete questionnaire data.

|  |  |  |  |  |  |  |  |  |  |  |  |  |
| --- | --- | --- | --- | --- | --- | --- | --- | --- | --- | --- | --- | --- |
|  | NO-KA group |  |  |  |  |  | KA group 2 years after surgery |  |  |  |  |  |
| | N (% of completer population) | Baseline<br>Mean (SD) | 12-week FU<br>Mean (SD) | 2-year FU<br>Mean (SD) | Δscore<sub>2y FU-baseline</sub><br>Mean diff (95% CI) | Δscore<sub>2y FU-12w FU</sub><br>Mean diff (95% CI) | N (% of full population) | Baseline<br>Mean (SD) | 12-week FU<br>Mean (SD) | 2-year PostOP<br>Mean (SD) | Δscore<sub>2y PostOp-baseline</sub><br>Mean diff (95% CI) | Δscore<sub>2y PostOp-12w FU</sub><br>Mean diff (95% CI) |
| OKS, points | 51 (76.1) | 26.9 (7.4) | 34.0 (8.6) | 31.3 (9.3) | 4.4 (1.9, 6.9)* | -2.7 (-4.8, -0.5) | 38 (76.0) | 25.1 (7.7) | 25.5 (7.5) | 37.9 (9.4) | 12.8 (9.9, 15.7)* | 12.4 (9.4, 15.4)* |
| KOOS |  |  |  |  |  |  |  |  |  |  |  |  |
| Pain, KOOS points | 51 (76.1) | 53.8 (16.8) | 69.6 (18.1) | 65.2 (21.2) | 11.4 (6.1, 16.7)* | -4.3 (-9.4, 0.7) | 38 (76.0) | 48.8 (16.5) | 49.9 (17.6) | 85.4 (19.7) | 36.7 (29.7, 43.7)* | 35.6 (28.5, 42.6)* |
| ADL, KOOS points | 50 (74.6) | 60.0 (18.9) | 74.5 (19.0) | 69.2 (20.9) | 9.1 (3.8, 14.4)* | -5.3 (-10.1, -0.5) | 38 (76.0) | 54.6 (17.1) | 57.3 (16.5) | 83.2 (21.1) | 28.6 (22.3, 34.9)* | 25.9 (20.0, 31.8)* |
| QoL, KOOS points | 51 (76.1) | 37.0 (17.2) | 51.1 (20.1) | 50.0 (23.2) | 13.0 (7.0, 19.0)* | -1.1 (-7.0, 4.8) | 37 (74.0) | 29.1 (15.9) | 32.8 (17.5) | 69.1 (26.6) | 40.0 (31.9, 48.2)* | 36.3 (28.4, 44.2)* |
| Symptoms, KOOS points | 51 (76.1) | 56.7 (19.7) | 72.1 (18.4) | 66.8 (22.2) | 10.1 (4.6, 15.6)* | -5.3 (-10.6, -0.1) | 38 (76.0) | 56.4 (20.0) | 58.0 (22.4) | 85.1 (18.0) | 28.7 (21.6, 35.8)* | 27.1 (20.0, 34.1)* |
| Sport, KOOS points | 50 (74.6) | 22.3 (21.1) | 39.0 (25.8) | 34.4 (28.3) | 12.0 (4.0, 20.1)* | -4.6 (-12.5, 3.3) | 35 (70.0) | 20.1 (18.8) | 23.9 (21.5) | 54.0 (28.3) | 33.9 (24.1, 43.6)* | 30.1 (21.9, 38.4)* |
| Note: Data is shown for participants with complete data from the 2-year follow-up questionnaire, which is why values and N differ from Table 2; “% of completer population”=percentage of the 117 participants who completed the 12-week QUADX-1 intervention. *Bonferroni-corrected P-values < 0.05 for within-group difference.<br>Abbreviations: ADL=Activities of Daily Living; 95% CI=95% confidence interval; Diff=Difference; FU=follow up; KA=knee arthroplasty; KOOS=Knee Osteoarthritis Outcome Score (0-100, best); N=number; OKS=Oxford Knee Score (0-48, best); PostOp=postoperative follow-up (mean 1032 days after surgery); for participants who underwent KA within 2 years of completing the 12-week QUADX-1 intervention, follow-up was scheduled 2 years after surgery; QoL=Quality of Life; SD=standard deviation; 12w FU=12-week follow-up assessment, 2y FU=2-year follow-up assessment. |  |  |  |  |  |  |  |  |  |  |  |  |

## DISCUSSION

The primary finding of this 2-year follow-up to the QUADX-1 trial was that more than half (57%) of the patients initially considered eligible for a KA—who completed a simple 12-week home-based exercise intervention—had still not received a KA at two years. Those who remained surgery-free at two years demonstrated less severe radiographic OA at baseline and reported greater improvements across outcome constructs, such as knee pain and functional performance, after the 12-week exercise intervention.

When compared with the existing literature, our main findings align with prior studies suggesting that non-surgical management, including exercise therapy, can lead to clinically meaningful symptom improvements and delay surgery for a significant proportion of patients with severe knee OA. For instance, Skou et al. (2018), Dell’Isola et al. (2021) and Dabare et al. (2017) demonstrated similar reductions in KA rates following more comprehensive and primarily supervised exercise interventions (6,8,9). A key distinction of this study is our use of a minimalistic intervention, insofar as we used a single knee-focused exercise performed at home after initial instruction. This contrasts with the more comprehensive, and in some cases supervised, non-surgical interventions used in those three trials (6,8,9), yet the QUADX-1 intervention achieved comparable long-term outcomes. Although not a randomized head-to-head comparison, it does suggest that the type of exercise and dose is of less importance for outcomes related to knee symptoms and the decision to have surgery or not. Given that so different exercise programs and delivery forms seem to achieve comparable outcomes, this raises the possibility that other factors, such as attention from health care professionals, may play a greater role. Attention (from health care professionals) has indeed been shown to yield comparable results to some types of exercise therapy in knee osteoarthritis (19).

Our findings also align with observations from other studies that baseline symptom severity (7) and radiographic OA severity (20) may be important in discussing possible progression to surgery when non-operative management is unsuccessful. Besides baseline radiographic OA grading, we found that the response to the 12-week exercise intervention seemed related to progression to surgery.

That is, compared with the KA group, the NO-KA group had greater—and often clinically relevant—improvements across outcomes, including self-reported knee pain, other symptoms and ADL-functioning (Table 2). These improvements were generally sustained at two years. In the subgroup with complete 2-year questionnaire data, 2-year OKS and KOOS values in the NO-KA group were descriptively lower than the corresponding postoperative values in the KA group (Table 3). This finding, although exploratory, is compatible with the notion that symptom response to QUADX-1 exercise helped identify which patients could continue without surgery and which patients were better served by KA. So, returning to the question stated in the introduction about potentially just kicking the can down the road, these data, although exploratory, suggest that for many patients “the can” can be kicked down the road for at least two years. The data also suggest that in a model of coordinated non-surgical and surgical care of patients with severe knee OA, the symptom response to 12 weeks of exercise may help inform the decision of whether to have surgery (21).

We recently advocated for a shift in the narrative for exercise-based prehabilitation when used in severe knee OA when a KA is being considered (21). In these cases, the focus is typically to use exercise-based prehabilitation to enhance post-operative recovery because it has been decided to have surgery, not to help inform the surgical decision. Rather than focusing solely on enhancing postoperative recovery, prehabilitation—or “pre-evaluation exercise” as we suggest (21)—can be framed as a tool for enhancing surgical decision-making. This aligns with the QUADX-1 trial model, where a simple home-based exercise intervention was intended to allow patients and clinicians to make more informed decisions about the need for surgery based on symptom changes. Importantly, this approach was also chosen to help support guideline recommendations and help bridge the gap between non-surgical and surgical care. Similar care pathways have subsequently been introduced in routine practice in the Capital Region in Denmark, and their long-term consequences are currently being evaluated prospectively at scale (22).

The mechanisms underpinning the observed differences in surgical outcomes likely relate to both symptom severity at baseline and the degree of symptom responsiveness to exercise. Prior studies have shown that early improvements in pain and function predict sustained avoidance of surgery (6,8,9) as outlined above. Additionally, reduced arthrogenic muscle inhibition (23) and psychosocial factors such as increased self-efficacy and perceived control (24) could potentially have contributed. However, despite large differences in exercise volume across QUADX-1 groups, changes in knee-extension strength were similar in response to the 12-week intervention (5), making differences in arthrogenic muscle inhibition less likely. The same can be said for psychological factors that may be the result of attention from health care professionals, as exposure to health care professionals was the same across QUADX-1 groups. However, increased self-efficacy and perceived control resulting from other sources than attention from health care professionals cannot be ruled out. A significant number of participants in the NO-KA group reported having used the exercise band after the 12-week exercise intervention ended. One could argue that it indicates some degree of self-efficacy and control. Finally, it is also plausible that patients who experienced greater early symptom relief had less central pain sensitization (25), which enabled longer-lasting benefits.

A key strength of this study is providing data to inform discussions on the long-term effects of—in this case—a minimal and very simple exercise intervention in patients with severe knee OA initially considered eligible for a KA. The simplicity of the exercise program makes it broadly accessible and likely cost-effective. As such, it may address common barriers to participation such as need for supervision, physical attendance, and financial costs. A limitation of this study is its exploratory nature, as the 2-year follow-up was not pre-specified.

## Conclusion

Two years after the initial 12-week QUADX-1 exercise intervention, more than half the cohort had not received a KA despite initially being considered eligible. Those who had not received a KA at two years had less severe radiographic OA at baseline, and they generally responded better to the 12-week exercise intervention two years earlier, compared to those who had.

## Data Availability

Public deposition of raw data points is not possible due to Denmarks national legislation (Data Protection Act Section 10 and Data Disclosure Proclamation Act) which outline that we can only transfer pseudonymized data to a publishing Journal or preprint server after the Data Protection Authorities approval (Data Protection Act Section 10, subsection 3, nr. 3.). Reviewers and others may obtain access to the data by request, and after the Danish Data Protection Agency has approved of the data transfer from the Capital Region to the Journal or repository entity. If others are to gain access to the pseudonymized data, the Journal or repository entity shall ensure that is an adequate legal basis to share the Capital Regions data and ensure that the data is only being processed for scientific research purposes.

## Acknowledgements

We thank research assistant Line Holst for help with trial management and outcomes assessments; the physiotherapists in the participating municipalities (Brøndby, Hvidovre and Copenhagen); and the orthopedic surgeons at the Department of Orthopedic Surgery, Copenhagen University Hospital, Hvidovre.

## Author contributions (QUADX-1 trial + follow-up)

Conceptualization: TB, KT, RSH, AT, BG, HGJL; Design and methodology: All authors; Experiments and data collection: RSH, BG; Data analysis: HGJL, BG, Data interpretation: All authors; Initial manuscript draft preparation: TB; Critical review and revision of subsequent manuscript drafts: All authors; All authors read and approved the final manuscript draft.

## Data sharing plan

Public deposition of raw data points is not possible due to Denmark’s national legislation (Data Protection Act § 10 and Data Disclosure Proclamation Act) which outline that we can only transfer pseudonymized data to a publishing Journal or preprint server after the Data Protection Authorities approval (Data Protection Act § 10, section 3, nr. 3.). Reviewers and others may obtain access to the data by request, and after the Danish Data Protection Agency has approved of the data transfer from the Capital Region to the Journal or repository entity. If others are to gain access to the pseudonymized data, the Journal or repository entity shall ensure that is an adequate legal basis to share the Capital Regions data and ensure that the data is only being processed for scientific research purposes.

